# Determinants of plasma levels of gcg and metabolic impact of glucagon receptor signalling – a UK Biobank study

**DOI:** 10.1101/2023.12.12.23299852

**Authors:** Marie Winther-Sørensen, Sara L. Garcia, Andreas Bartholdy, Maud Eline Ottenheijm, Karina Banasik, Søren Brunak, Charlotte M. Sørensen, Lise Lotte Gluud, Filip K. Knop, Jens Juul Holst, Mette M. Rosenkilde, Majken K. Jensen, Nicolai J. Wewer Albrechtsen

**Author notes:** Correspondence to Staff Specialist, Associate Professor, MD, PhD Nicolai J. Wewer Albrechtsen, Copenhagen University Hospital – Bispebjerg and Frederiksberg, Copenhagen, Denmark & Novo Nordisk Foundation Center for Protein Research, Faculty of Health and Medical Sciences, University of Copenhagen, Copenhagen, Denmark.

## Abstract

**Aims/hypotheses:** Glucagon and Glucagon-like peptide-1 (GLP-1) are derived from the same precursor; proglucagon (gcg), and dual agonists of their receptors are currently explored for the treatment of obesity and steatotic liver disease. Elevated levels of endogenous glucagon (hyperglucagonaemia) have been linked with hyperglycaemia in individuals with type 2 diabetes but are also observed in individuals with obesity and metabolic dysfunction-associated steatotic liver disease (MASLD). It is unknown whether type 2 diabetes, obesity or MASLD causes hyperglucagonaemia or vice versa. We investigated potential determinants of plasma gcg and associations of glucagon receptor signalling with metabolic diseases based on data from the UK Biobank.

**Methods:** We used exome sequencing data from the UK Biobank for ∼410,000 Caucasians to identify glucagon receptor variants and grouped them based on their known or predicted signalling. Plasma levels of gcg estimated using Olink technology was available for a subset of the cohort (∼40,000). We determined associations between glucagon receptor variants and gcg with BMI, type 2 diabetes, and liver fat (quantified by liver MRI) and performed survival analyses to investigate if elevated gcg predicts type 2 diabetes development.

**Results:** Obesity, MASLD, and type 2 diabetes independently associated with elevated plasma levels of gcg. Baseline gcg levels were statistically significantly associated with the risk of type 2 diabetes development over a 14-year follow-up period (hazard ratio = 1.13; 95% confidence interval (CI) = 1.09, 1.17, p < 0.0001). This association was of the same magnitude across strata of BMI. Carriers of glucagon receptor variants with reduced cAMP signalling had elevated levels of gcg (β = 0.847; CI = 0.04, 1.66; p = 0.04), and carriers of variants with a predicted frameshift mutation had significantly higher levels of liver fat compared to wild-type controls (β = 0.504; CI = 0.03, 0.98; p = 0.04).

**Conclusions/interpretation:** Our findings support that glucagon receptor signalling is involved in MASLD and type 2 diabetes, and that plasma levels of gcg are determined by genetic variation in the glucagon receptor, obesity, type 2 diabetes, and MASLD. Determining the molecular signalling pathways downstream of glucagon receptor activation may guide the development of biased GLP-1/glucagon co-agonist with improved metabolic benefits.

**Research in context:** What is already known about this subject?

- Glucagon contributes to fasting hyperglycaemia in type 2 diabetes
- Hyperglucagonemia is often observed in metabolic dysfunction-associated steatotic liver disease (MASLD), obesity and type 2 diabetes
- Glucagon/GLP-1 co-agonists have superior metabolic benefits compared to monoagonists

What is the key question?

What are key determinants of plasma proglucagon (gcg) and is elevated plasma gcg a cause or consequence (or both) of type 2 diabetes?

What are the new findings?

- Plasma levels of gcg are increased in type 2 diabetes, MASLD and obesity independently of each other
- Increased plasma gcg associates with higher risk of type 2 diabetes development
- Glucagon signalling associates with hepatic fat

How might this impact on clinical practice in the foreseeable future?

- Biased glucagon receptor-regulating agents may be beneficial in the treatment of obesity and MASLD.

## 1. Introduction

The proglucagon gene (*GCG*) encodes several metabolically important hormones including glucagon-like peptide-1 (GLP-1) and glucagon with impact on glucose control, food intake, and hepatic protein- and lipid metabolism [1]. Recently, co-agonists of these two hormones have been developed and are tested in clinical trials for the treatment of obesity and metabolic dysfunction-associated steatotic liver disease (MASLD) [2, 3]. Increased plasma levels of glucagon (hyperglucagonemia) are associated with fasting hyperglycaemia in patients with type 2 diabetes but are also observed in individuals with obesity and/or MASLD [4–6]. Glucagon binds and acts via the glucagon receptor, belonging to class B1 of the superfamily of G protein-coupled receptors signalling through Gα_s_ (stimulating the adenylate cyclase/cAMP/protein kinase A pathway) and Gαq (signalling through the phospholipase C/inositol-phosphate (IP3)/calcium/calmodulin pathway). Like other class B1 receptors, the glucagon receptor recruits β-arrestin, which sterically alters the binding between the receptor and the G protein and regulates internalization [7]. The molecular pharmacological phenotype of 38 missense variants of the glucagon receptor were recently described at the level of cAMP signalling and β-arrestin recruitment [8], whereas similar systematic investigations at the level of the PLC/IP3 pathway are still lacking.

During the last decade, the impact of glucagon signalling on hepatic amino acid catabolism has gained increasing interest due to its potential clinical implications. Equally important has been the recognition of the role of amino acids as determinants of hyperglucagonemia in patients with MASLD. We and others have shown that MASLD is associated with hepatic glucagon resistance resulting in hyperaminoacidemia and compensatory hyperglucagonemia, increasing hepatic glucose production [9, 10]. This feedback system, known as the liver-alpha cell axis, may contribute to the increased risk of type 2 diabetes observed in obese individuals with MASLD. The potent impact of glucagon on amino acid levels has also prompted the use of circulating amino acids (alanine in particular) as markers for drug engagement in studies investigating the clinical utility of glucagon receptor co-agonism [11].

An important gap in the understanding of the pathophysiological role of glucagon in metabolic diseases lies in elucidating whether increased plasma levels of glucagon result from a) obesity, b) MASLD, c) type 2 diabetes, or d) a combination of these. A key question in this context is whether hyperglucagonemia is merely an epiphenomenon of dysmetabolic conditions or a direct contributor to the development of type2 diabetes. Elucidation of this has been challenged due to lack of sufficient matching of body weight, age, sex, MASLD status, and kidney function, in particular because of limited sample sizes across reported clinical studies.

To determine glucagon’s role in type 2 diabetes, and whether hyperglucagonemia exists independently of obesity, MASLD and type 2 diabetes, we analysed data from the UK Biobank including data from more than 500,000 individuals. The dataset included plasma gcg on ∼40,000 individuals, ∼15 years follow up data on incident type 2 diabetes development, amino acid quantification for ∼230,000 individuals, liver fat quantification for ∼35,000 individuals, and exome sequencing allowing investigations of the potential impact of glucagon receptor variants on clinical features.

## 2. Materials and Methods

### 2.1. The UK Biobank

The UK Biobank (UKB) is a large prospective research resource including half a million participants aged 40 to 69 at the time of inclusion from the United Kingdom. The biobank encompasses genetic, lifestyle, and health data derived from various sources such as questionnaires, physical assessments, biological specimens, imaging, and the continual monitoring of health-related outcomes, as described in detail previously [12]. Participants who withdrew from the biobank were excluded from all analyses (n = 179, updated 14.11.2023). We excluded non-white individuals and outliers for sex chromosome aneuploidy. A list of field names used in the study is available in ESM Table 1.

### 2.2. Proteomics data processing

Proteomics data was accessed through the DNA nexus platform and processed locally. The data is available as NPX units (an arbitrary and normalized unit in Log2 scale). Processing of the raw data and normalization has been described elsewhere [13]. Overall, there was 2.9% missing data. We filtered out individuals and proteins with >10% missing data (4 proteins and 3298 individuals were excluded). The remaining 1.1% missing data was imputed by MinProb. The resulting Olink sub-cohort consisted of 40,164 individuals and 1460 proteins. Differential expression analysis was performed with the Limma package (v. 3.56.2) [14].

### 2.3. Exome Sequencing Analysis and Variant Annotation

We analysed the whole-exome sequencing of 469,914 individuals from the UKB [12]. The UKB whole-exome sequencing data was reference-aligned with the Original Quality Functional Equivalent protocol previously described [15]. This protocol uses BWA-MEM [16] to map all the reads to the human reference genome GRCh38 [17]. Variant call was performed using DeepVariant [18]. We filtered the Genomic VCF (gVCF) files for each sample, restricted to the location of the glucagon receptor at chr17: 81,804,150 to 81,814,008 forward strand. The analyses were conducted on the Research Analysis Platform (https://ukbiobank.dnanexus.com). A cloud worker with 36 CPU threads was chosen to run 36 bcftools (swiss army knife) parallel jobs and automatically launch new jobs when the previous jobs had finished. The output csv files were subsequently merged using Python. The scripts used to filter the gVCF files from UKB and merge the output csv files are available at https://github.com/nicwin98/UK-Biobank-GCG.

We filtered for genotype quality (GQ) < 20, depth (DP) < 10, and allele balance (AB, for the minor allele) < 0.2. The genetic variants were annotated for their sequence effect with opencravat.org [19]. We created a variant group “Frameshift” for predicted loss-of-function alleles. This group included the sequence ontologies frameshift elongation, frameshift truncation, in-frame deletion, in-frame insertion, start lost, and stop gained (see ESM Table 2). Missense variants were denoted as the reference amino acid (1 letter code), the amino acid position, followed by the alternative amino acid. Missense variants were subsequently categorized as G40S heterozygotes and G40S homozygotes (pooled in the proteomics sub-cohort), or as cAMP loss-of-function based on a previous study reporting the molecular phenotype of 38 missense variants [8].

### 2.4. Incident type 2 diabetes and survival analyses

We defined type 2 diabetes based on hospital diagnoses encoded as E11 or E14 in the ICD-10 classification system. We excluded individuals with a diagnosis of type 1 diabetes (E10). Prevalent cases were defined as 1) probable and possible type 2 diabetes based on the Eastwood algorithm [20], 2) with a baseline HbA1c greater than 48 mmol/mol (a recommended cutoff point for diagnosing type 2 diabetes) [21], or 3) a diagnosis before or within 6 months after the enrolment visit. A list of fields used for the definition of diseases is available in ESM Table 3. After exclusion of 1833 prevalent cases, a total of 1562 developed incident type 2 diabetes during follow-up (median follow-up time: 14.75 years).

Risk time (in months) was defined from date of baseline examination (between 2006 and 2010) where the blood sample for proteomics analysis was obtained, to the last updated version of the hospital register (3^rd^ of October 2021), the date of type 2 diabetes diagnosis, death, or loss to follow-up, whichever occurred first. Data on loss to follow-up was last updated in May 2017, and censored individuals were included in the analysis up to the point of censoring.

For the Kaplan-Meier survival analysis, the cohort was stratified into tertiles based on their plasma gcg levels at the baseline visit. Pairwise comparison using Log-Rank test with Bonferroni correction for multiple testing was used to compare survival curves between the three subgroups. Using Cox proportional hazard regression with adjustment for age and sex, and we explored BMI (continuously) as a potential intermediate. However, due to lacking model fit our final analysis instead stratified on BMI categories (BMI < 25 kg/m^2^; 25 kg/m^2^ ≥ BMI > 30 kg/m^2^; BMI ≥ 30 kg/m^2^) and addressed whether the association between gcg and incident type 2 diabetes differed across BMI categories. Proportional Hazards and linearity assumptions for all covariates were assessed with Schonfeld and Martingale residual plots, respectively. Survival analyses were done using the Survival package for R (v3.5-4; Therneau, 2020) [22].

### 2.5. Statistical methods

For association analyses, we used the following variables: Age when attending the assessment centre; sex; BMI; baseline type 2 diabetes was defined as probable and possible type 2 diabetes based on the Eastwood algorithm [20]; liver fat (quantified by MRI-PDFF at the second repeat visit) [23]; weekly alcohol; glucose; Hba1c; amino acids. BMI was used continuously or as a binary variable (non-obese, BMI < 25 kg/m^2^; obese, BMI > 30 kg/m^2^). Liver fat was used as a continuous variable or a binary variable to define MASLD (non-MASLD, < 5.5%, MASLD ≥ 5.5%), in both cases excluding individuals with a weekly alcohol intake above 17.5 units for women and 26.25 units for men. The gcg-alanine index was calculated as the product of gcg and alanine.

For linear models, increments of independent variables were set to 5 units for BMI, 5% for liver fat, 5 years for age, and 5 mmol/L for serum creatinine. Glucose, Hba1c, and amino acids were normalized to the standard deviation (SD) of each variable. Glucagon receptor variant groups were tested for association to binary traits with logistic regression with Firth-correction. R version 4.3.0 was used for all analyses.

## 3. Results

### 3.1. Plasma gcg is elevated in obesity, type 2 diabetes, and MASLD and associates with increased risk of incident type 2 diabetes

Plasma gcg was measured as a part of the proteomics analysis in a sub-cohort that matched the UKB cohort on age, sex, and recruitment centre (ESM Table 4) [13]. We investigated whether plasma gcg was associated with BMI, type 2 diabetes, and MASLD in the UK Biobank. Plasma gcg was available as NPX units (an arbitrary and normalized unit in Log2 scale). Plasma gcg was increased in individuals with obesity (Log2 fold change (FC) = 0.53, p < 0.0001) (Fig. 1a), type 2 diabetes (Log2 FC = 1.10, p < 0.0001) (Fig. 1b), and MASLD (Log2 FC = 0.36, p < 0.0001) (Fig. 1c).

We performed multiple linear regression models to assess the association between plasma gcg as independent variable and type 2 diabetes, BMI, and liver fat as dependent variables. Model 1 was adjusted for age, sex, fasting time, and plasma creatinine. All variables significantly associated with higher plasma gcg (Fig. 1d). To investigate whether these metabolic diseases independently associated with higher gcg levels, we adjusted the linear models for each of the other variables. Importantly, plasma gcg remained positively associated with type 2 diabetes (p < 0.0001), BMI (p < 0.001) and liver fat (p < 0.01) (Fig. 1d) suggesting that each of these metabolic disorders independently associate with elevations in plasma gcg. This was confirmed in an additional analysis with a tenfold increase in the sample size for type 2 diabetes and BMI specifically (Fig. 1e).

**Fig. 1:**
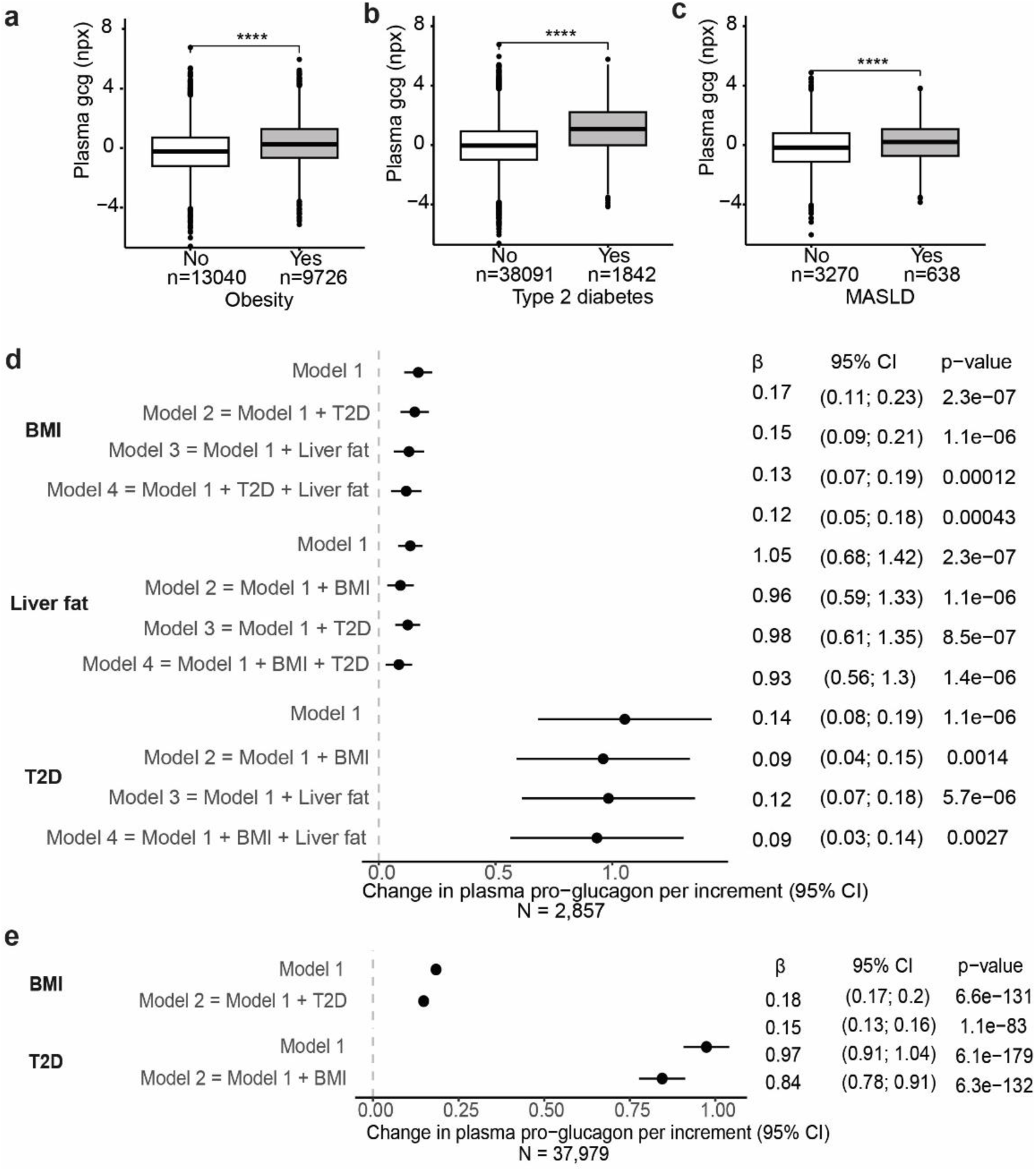
Plasma gcg is elevated in obesity, type 2 diabetes, and MASLD. Plasma gcg in (a) lean individuals (BMI < 25) and individuals with obesity (BMI > 30), (b) individuals with and without type 2 diabetes, and (c) individuals with MASLD. The number of individuals in each group is shown. Data is shown as boxplots with quartiles and analysed by unpaired t-test. ***P<0.001, ****P < 0.0001. (d) Multiple linear regression analyses were performed with plasma gcg from Olink proteomics as the dependent variable and T2D, BMI, % liver fat (PDFF) as independent variables. Increments were set to 5 units for BMI and 5% for liver fat. Model 1 for each variable included adjustment for age, sex, fasting time, and plasma creatinine. Additional co-factors in each of the remaining models are indicated at the figure. (e) Multiple linear regression analyses with plasma gcg as the dependent variable and T2D and BMI as independent variables. Increments were set to 5 kg/m^2^ for BMI. Model 1 for each variable included adjustment for age, sex, fasting time, and plasma creatinine. Additional co-factors in each of the remaining models are indicated in the figure. CI, 95% confidence intervals; T2D, type 2 diabetes.

To investigate if high plasma levels of gcg associate with an increased risk of developing type 2 diabetes, we first performed a Kaplan-Meier survival analysis stratified on tertiles of plasma gcg at baseline (Tertile 1: mean gcg (NPX): -1.50, n=13385; Tertile 2: mean gcg: -0.005, n=13386; Tertile 3: mean gcg: 1.62, n=13385). The median follow-up time was 14.75 years, and the number of incident type 2 diabetes cases included in the models was 1551 (893 men and 658 women). The risk of incident type 2 diabetes increased stepwise from low to medium levels (p < 0.0001) and from medium to high levels (p < 0.0001) (Fig. 2a). Secondly, we applied a Cox proportional hazard regression analysis to evaluate the impact of baseline gcg levels of incident type 2 diabetes with adjustment for age and sex. Gcg statistically significantly associated with the risk of type 2 diabetes development (HR: 1.13; CI = 1.09. 1.17, p < 0.0001). We also stratified our model on BMI, and the association between gcg and incident type 2 diabetes was of similar magnitude across BMI categories (Fig. 2b).

**Figure 2:**
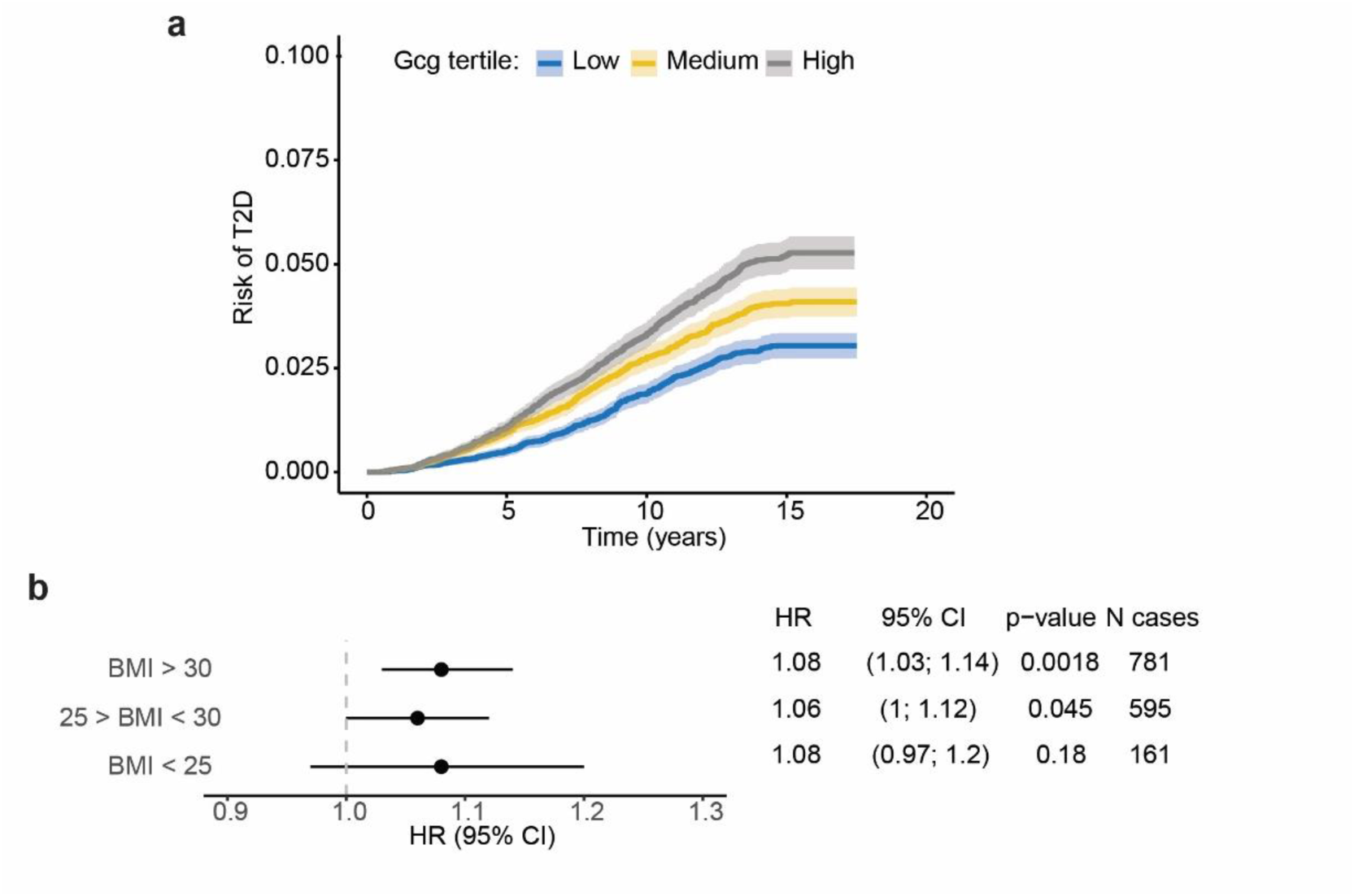
**(a)** Kaplan-Meiner survival curves for incident type 2 diabetes during the follow-up period, differentiated by tertiles of baseline gcg levels. The shaded areas represent 95% confidence intervals. Tertile 1: n = 13,385, mean gcg = -1.50 NPX; Tertile 2: n = 13,386, mean gcg = -0.005 NPX; Tertile 3: n = 13,385, mean gcg = 1.62 NPX. The subgroups were statistically compared using pairwise Log-Rank test with Bonferroni correction. (b) Results from Cox proportional hazard regression analysis. The model was stratified on BMI categories: normal-weight, BMI < 25 kg/m^2^; overweight, 25 kg/m^2^ ≥ BMI > 30 kg/m^2^; obese, BMI ≥ 30 kg/m^2^. Gcg (NPX) was the independent variable and age (5-year increment) and sex were included as covariates. HR, hazard ratio; CI, 95% confidence interval.

### 3.2. Association between plasma gcg and circulating metabolites

We tested the impact of selected confounders on plasma levels of gcg using linear models. Gcg levels were higher in males than females and increased with age (Fig. 3a). Plasma creatinine was included as an estimate of renal function, as circulating products of gcg are cleared in the kidneys. An increase in plasma creatinine, suggestive of reduced renal clearance, was associated with an increase in gcg (Fig. 3a).

Next, we used multiple linear regression to evaluate the association between plasma gcg and amino acids and glucose, adjusting for BMI and the confounders in Fig. 3a. Plasma gcg was positively associated with tyrosine, phenylalanine, alanine and histidine as well as the branched-chain amino acids valine, leucine, and isoleucine. Interestingly, glutamine and glycine were not associated with gcg levels (Fig. 3b). Plasma glucose and Hba1c were also positively associated with gcg levels (Fig. 3b).

The glucagon-alanine index has been proposed as a plasma marker for glucagon resistance [24–28]. It is associated with hepatic steatosis [24], and decreases upon weight loss [25]. The gcg-alanine index (the product of gcg and alanine) correlated with liver fat (β = 0.15, p < 0.001) (Fig. 3c).

**Fig. 3:**
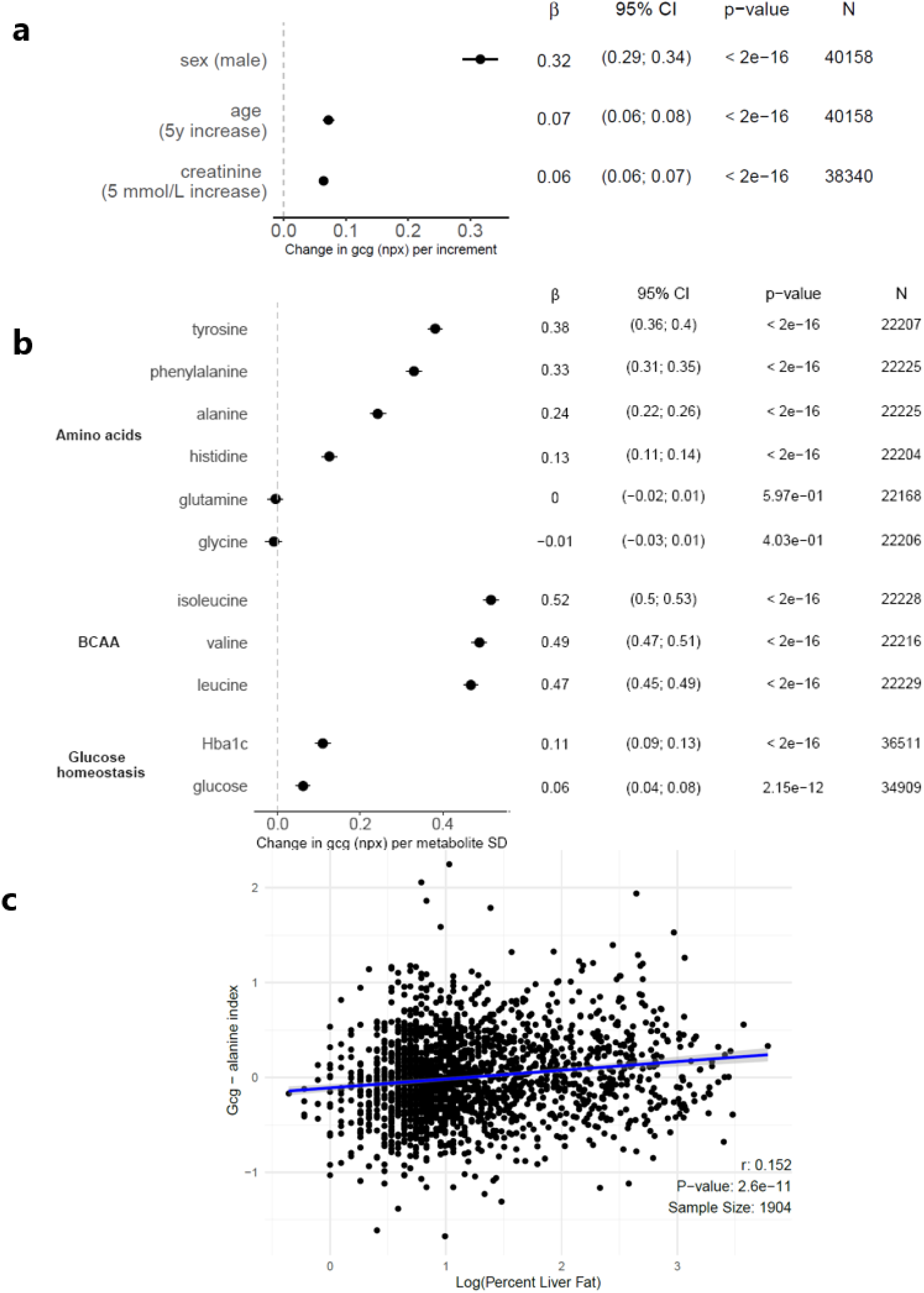
Effects of circulating metabolites on plasma gcg levels. (a) Simple linear regression analyses were performed with plasma gcg in NPX units as the dependent variable and the indicates confounders as independent variables. Increments were set to 5 years for age and 5 mmol/L for creatinine. (b) Multiple linear regression analyses were adjusted for BMI, sex, age, creatinine, and fasting time. The x-axis is the effect size in standard deviations (SD) of the metabolite. P-values were adjusted using FDR correction. (c) The gcg-alanine index (the product of gcg (NPX) and alanine (mmol/L)) plotted against percent liver fat (log scale). Correlation was tested using Pearson correlation test. r, correlation coefficient.

### 3.3. Associations between gcg and T2D and BMI may depend on genetic variation of the glucagon receptor

To investigate the effect of genetic variation in the glucagon receptor on the same metabolic outcomes as above, we identified individuals with genetic variants of the glucagon receptor from whole-exome sequencing data. We grouped the glucagon receptor variants the following way: 1) the single nucleotide polymorphism G40S, previously associated with non-insulin dependent diabetes, hypertension, and adiposity [29–31], but normal cAMP signalling and reduced β-arrestin signalling [8], 2) the missense variants V368M, R378C, R225H, R308W, and D63 were grouped as “cAMP loss-of-function” (LoF) based on previous research [8], and 3) variants annotated as frameshift or stop-codon introduced were grouped as “Frameshift” variants (ESM Table 2). Fig. 4a-c outlines the number of individuals in each group in the UK Biobank cohort and the sub-cohort included in the proteomics analysis.

Although only 11 individuals heterozygous for a cAMP LoF variant were included, we observed a statistically significant elevation in gcg compared to the wildtype reference group (β = 0.847; CI = 0.04, 1.66; P = 0.04) (Fig. 4d). G40S or frameshift variants were not associated with gcg levels, implying that cAMP rather than β-arrestin signalling may be involved in metabolic processes directly or indirectly regulating gcg levels.

In the frameshift variant group, liver fat was significantly increased compared to the wild-type reference group (β = 0.504; CI = 0.03, 0.98; p = 0.04) (Fig. 4e). None of the glucagon receptor variant groups were associated with BMI as a continuous or binary trait. We observed no difference in the prevalence of type 2 diabetes between G40S and controls (Fig. 4f). The sample size of type 2 diabetes was inadequate for cAMP LoF and pLoF variant groups.

To test if the association between plasma gcg and type 2 diabetes and BMI, respectively, was dependent on the glucagon receptor genotype, we performed logistic and linear models stratified on the variant groups and with inclusion of the interaction term gcg*genotype. Interestingly, although plasma gcg associated with type 2 diabetes in the whole cohort (Fig. 1d, 1e), this was not the case in individuals with G40S (p = 0.1) (Table 1). The interaction term between G40S and gcg was borderline significant (p = 0.07). Oppositely, the effect of plasma gcg on BMI was larger in carriers of the G40S variants compared to wildtype controls (0.679±0.126 vs. 0.327±0.017, p < 0.05) (Table 1). The effects of plasma gcg on BMI in carriers of cAMP LoF and truncated variants were not different from individuals with wildtype receptors, however, the results are limited by the low sample size (11 and 24, respectively) (Table 1).

**Fig. 4:**
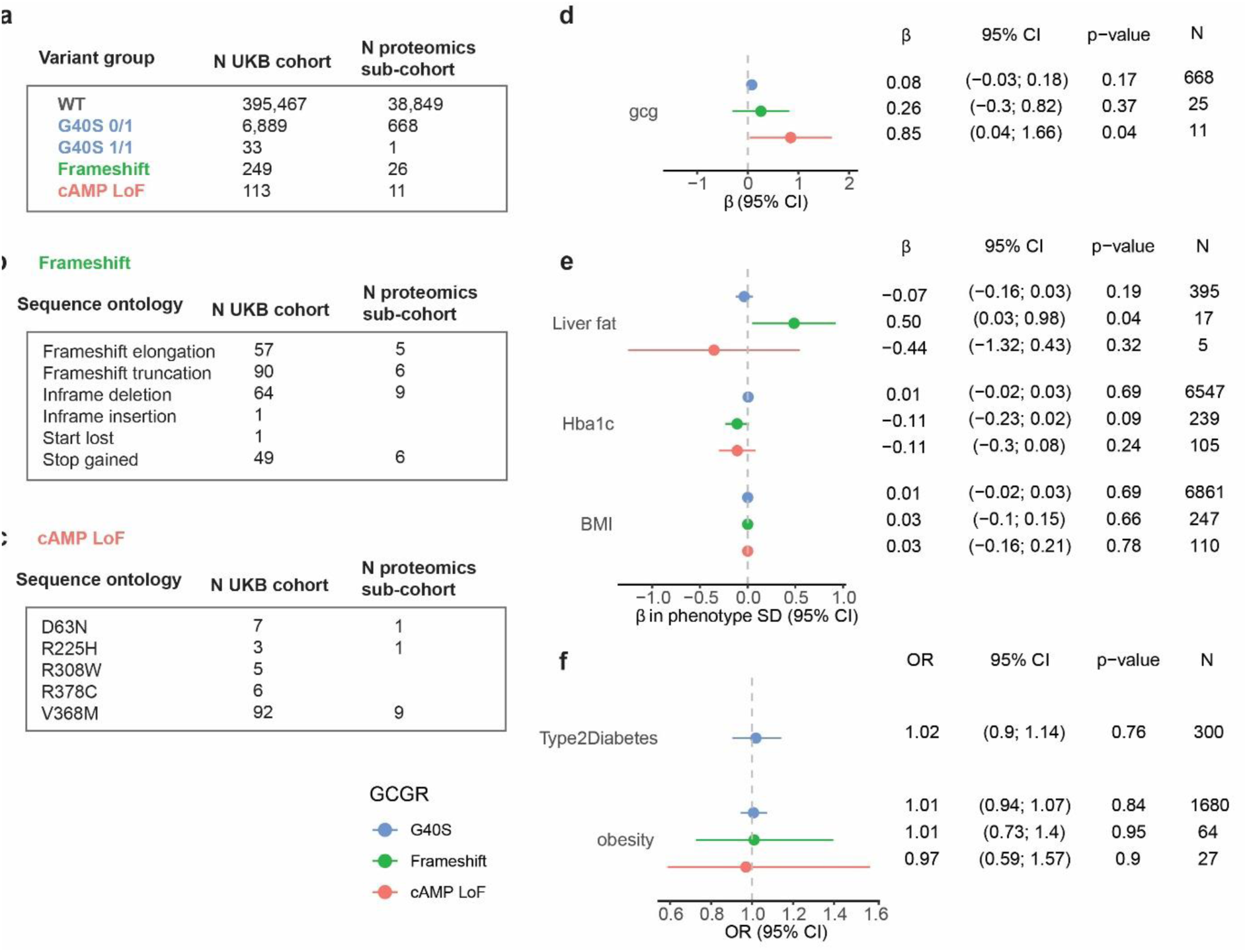
Association of loss-of-function variants with plasma gcg and metabolic traits. (a) Number of individuals within each glucagon receptor variant group in the UKB cohort and the proteomics sub-cohort. (b) The Frameshift variant group divided into the sequence ontology terms included in the group. (c) The cAMP LoF variant group divided according to the missense mutations included in the group. (d) Variant groups were tested for association to plasma gcg in a multiple linear model adjusted for age, sex, BMI, fasting time, and plasma creatinine. G40S heterozygous and homozygous were pooled. (e) Variant groups were tested for associations with quantitative traits in multiple linear models adjusted for age and sex. The x-axis is the effect size (β) in standard deviations (SD) of the phenotype. (f) Variant groups were tested for association to binary traits with logistic regression with Firth-correction adjusted for age and sex. Obesity was defined as BMI > 30 and controls as BMI < 25. N indicates the number of cases in each variant group. d, e, f: Blue, G40S; Green, frameshift variants; Red, cAMP loss-of-function variants. G40S was included as a numeric predictor (1, heterozygous; 2, homozygous). WT, wildtype; CI, confidence interval; OR, odds ratio.

**Table 1.** Interaction between plasma gcg and genotype on type 2 diabetes and BMI. For type 2 diabetes, model 1 was a logistic model with type 2 diabetes as the dependent variable and gcg as independent variable adjusted for age, sex, fasting time, creatinine, and BMI, and stratified on the genotype G40S. Model 2 included the interaction between gcg and G40S. For BMI, model 1 was a linear model with BMI as the dependent variable and gcg as independent adjusted for age, sex, fasting time, creatinine, and type 2 diabetes, and stratified on the glucagon receptor variant groups. Model 2 for each variant group included the interaction between gcg and variant group. WT, wildtype; T2D, type 2 diabetes, CI, confidence interval.

| Type 2 diabetes |  |  |  |  |  |  |
| --- | --- | --- | --- | --- | --- | --- |
| Genotype | N controls | N Type 2 diabetes | Effect of plasma gcg |  |  | Interaction between plasma gcg and genotype |
| | | | $\beta$ | CI | p-value | p-value |
| WT | 36819 | 776 | 0.464 | (0.43, 0.5) | < 0.0001 |  |
| G40S | 630 | 39 | 0.207 | (-0.04, 0.46) | 0.107 | 0.073 |
| BMI |  |  |  |  |  |  |
| Genotype | N |  | Effect of plasma gcg |  |  | Interaction between plasma gcg and genotype |
| | | | $\beta$ | CI | p-value | p-value |
| WT | 38662 |  | 0.327 | (0.29, 0.36) | < 0.0001 |  |
| G40S | 669 |  | 0.679 | (0.43, 0.93) | < 0.0001 | 0.013 |
| cAMP LoF | 11 |  | 2.984 | (-4.65, 10.62) | 0.339 | 0.141 |
| Frameshift | 24 |  | -0.1514 | (-1.85, 1.55) | 0.853 | 0.389 |

### 3.4. Circulating amino acids and proteins are not altered by loss-of-function glucagon receptor variants

Plasma levels of the individual and the sum of amino acids were not significantly different in individuals with the glucagon receptor variant groups compared to wildtype controls (table 2). However, a numeric increase (p = 0.10) in plasma alanine was observed in cAMP LoF in line with prior studies linking glucagon resistance to an increased glucagon-alanine index.

**Table 2:** Association between glucagon receptor variant groups and plasma amino acids. The associations were tested in linear models with the genotype as predictor and age, sex, BMI, and fasting time as covariates. β is given in amino acid SD. β, effect size; CI, 95% confidence interval.

|  | cAMP LoF |  |  |  | G40S |  |  |  | Frameshift |  |  |  | SD |
| --- | --- | --- | --- | --- | --- | --- | --- | --- | --- | --- | --- | --- | --- |
| | $\beta$ | CI | p-value | N | $\beta$ | CI | p-value | N | $\beta$ | CI | p-value | N | |
| Alanine | 0.20 | (-0.04; 0.45) | 0.10 | 63 | -0.01 | (-0.04; 0.02) | 0.66 | 3868 | -0.06 | (-0.22; 0.1) | 0.44 | 151 | 0.08 |
| Glutamine | 0.04 | (-0.2; 0.29) | 0.72 | 63 | 0.03 | (-0.01; 0.06) | 0.10 | 3863 | 0.11 | (-0.05; 0.26) | 0.18 | 151 | 0.08 |
| Glycine | -0.20 | (-0.43; 0.03) | 0.09 | 63 | 0.02 | (-0.01; 0.05) | 0.12 | 3860 | -0.06 | (-0.21; 0.09) | 0.42 | 151 | 0.07 |
| Histidine | -0.03 | (-0.28; 0.21) | 0.79 | 63 | -0.02 | (-0.05; 0.01) | 0.26 | 3861 | 0.08 | (-0.08; 0.24) | 0.33 | 151 | 0.01 |
| Isoleucine | -0.02 | (-0.26; 0.22) | 0.88 | 63 | 0.01 | (-0.02; 0.04) | 0.55 | 3869 | 0.02 | (-0.13; 0.18) | 0.76 | 151 | 0.02 |
| Leucine | -0.03 | (-0.26; 0.21) | 0.83 | 63 | 0.00 | (-0.03; 0.03) | 0.99 | 3869 | 0.03 | (-0.12; 0.18) | 0.68 | 151 | 0.03 |
| Valine | -0.13 | (-0.36; 0.1) | 0.28 | 63 | 0.00 | (-0.03; 0.03) | 0.93 | 3866 | 0.01 | (-0.14; 0.16) | 0.88 | 151 | 0.04 |
| Phenylalanine | -0.12 | (-0.36; 0.12) | 0.34 | 63 | 0.03 | (0; 0.06) | 0.08 | 3867 | -0.06 | (-0.22; 0.09) | 0.42 | 151 | 0.01 |
| Tyrosine | -0.03 | (-0.27; 0.21) | 0.79 | 63 | 0.02 | (-0.01; 0.05) | 0.18 | 3863 | -0.02 | (-0.18; 0.13) | 0.77 | 151 | 0.02 |
| Total AA | -0.01 | (-0.26; 0.23) | 0.92 | 63 | 0.02 | (-0.01; 0.05) | 0.27 | 3845 | 0.01 | (-0.15; 0.17) | 0.94 | 151 | 0.21 |

We next performed differential expression analysis on the proteomics dataset (∼1,500 proteins measured in each sample) to identify plasma proteins potentially regulated by glucagon receptor signalling. After correction for multiple testing, no proteins reached statistical significance. A list of the top 10 up- and downregulated proteins for each variant group is provided in Table 3.

**Table 3:**
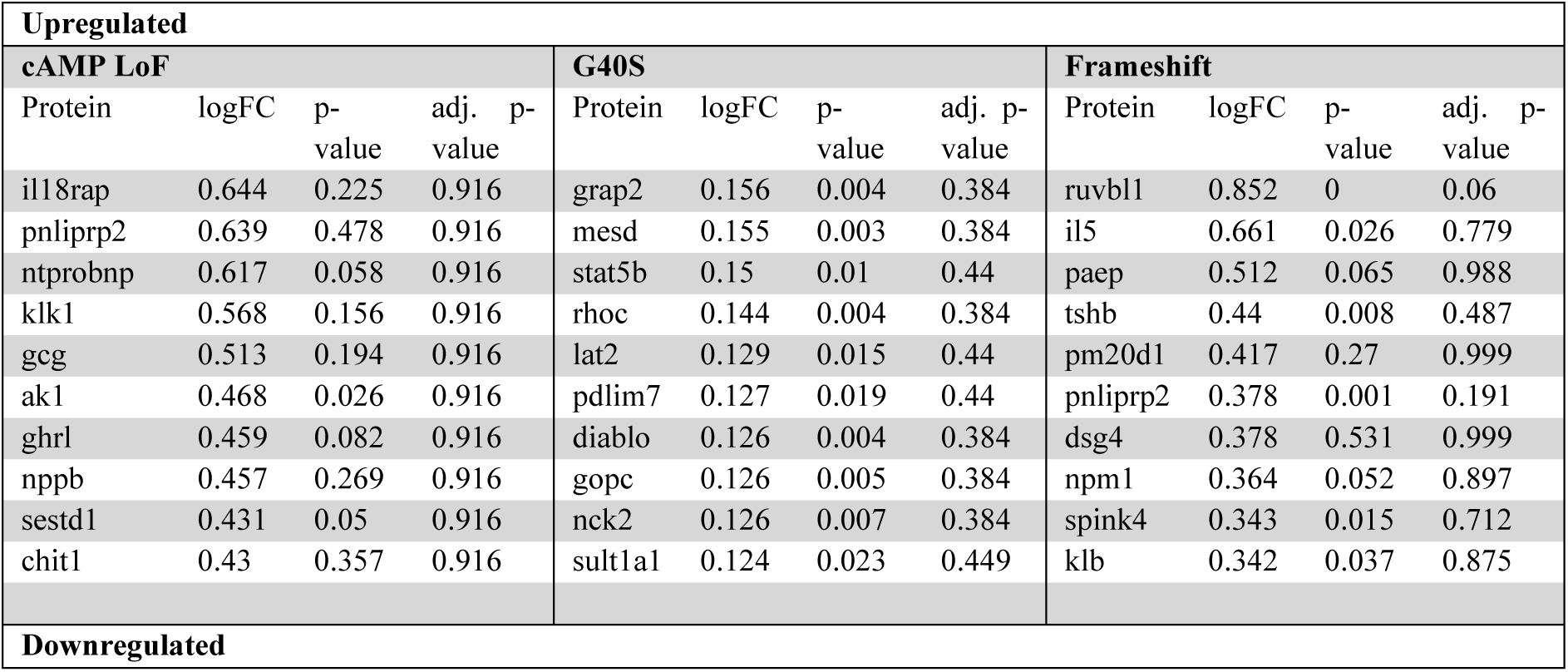
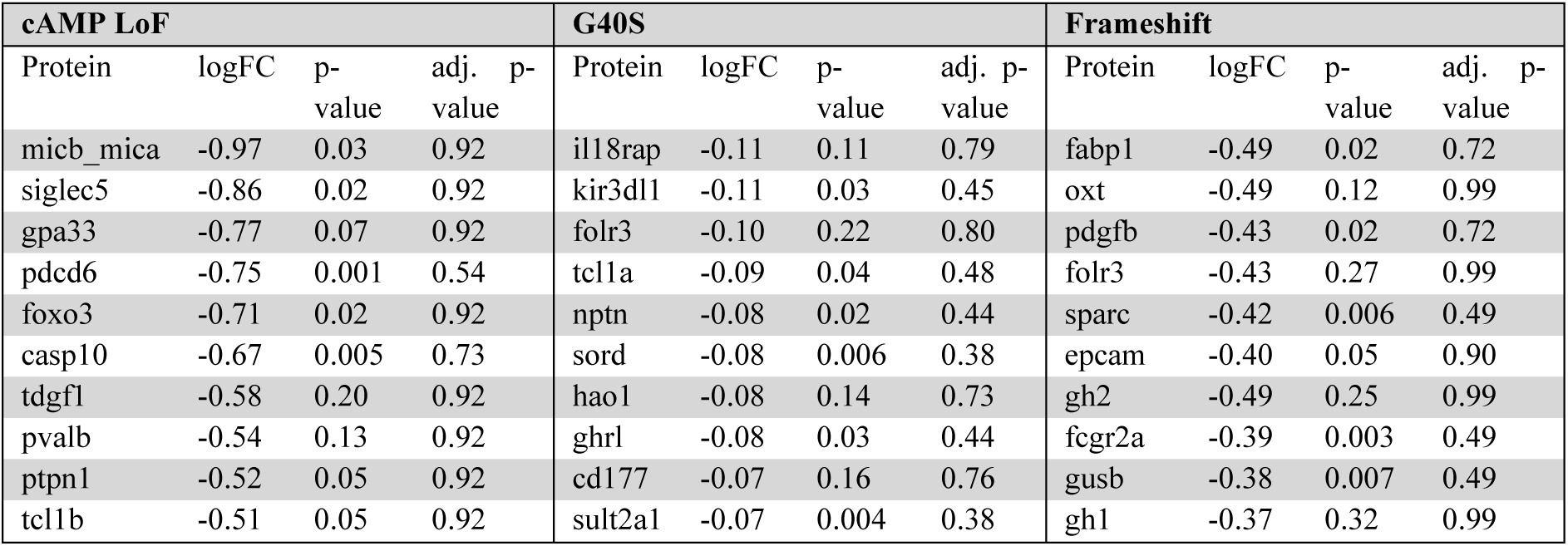
Top 10 up- and downregulated proteins in glucagon receptor variant groups. Differential expression analysis was performed for each of the variant groups. p-values were adjusted by FDR for multiple testing. logFC, log fold change.

## 4. Discussion

Using UKB data encompassing multiple omics data, MRI imaging and hospital registers, we here demonstrated that increased plasma levels of gcg are independently associated with obesity, MASLD, and type 2 diabetes. In addition, gcg levels were significant predictors of the risk of type 2 diabetes development over a 14-year follow-up period. Although causality cannot be drawn, our data support that increased glucagon directly contributes to development of type 2 diabetes [32].

One of the two antibodies in the Olink gcg assay binds within the first 100 amino acids of proglucagon. The assay may therefore measure both proglucagon products – namely glucagon and GLP-1. However, our results point towards glucagon as the measured peptide: Firstly, we observed that gcg was elevated in individuals with type 2 diabetes (Fig. 1b). Numerous studies have described that GLP-1 is reduced, and glucagon increased, in type 2 diabetes [33, 34]. Secondly, gcg was positively associated with alanine (Fig. 3b). Alanine was not correlated with plasma GLP-1 levels after a protein-rich intake in women with obesity [35] and did not stimulate GLP-1 secretion in vitro [36]. On the contrary, alanine is known to correlate positively with glucagon levels in the fasting and postprandial states [24, 37], and to strongly stimulate glucagon secretion across species [38–40]. Thirdly, the gcg-alanine index significantly correlated with liver fat. Multiple studies have shown that the glucagon-alanine index is correlated with liver fat even below the level of steatosis [26, 27, 41]. Lastly, we observed a significant increase in plasma gcg in carriers of cAMP LoF variants of the glucagon receptor (Fig. 4d). Since glucagon resistance in MAFLD results in a compensatory increase in glucagon levels [27, 41], similar mechanisms likely underlie the finding of increased gcg in individuals with reduced glucagon receptor activity at the level of cAMP signalling. The assumption that gcg represents glucagon rather than GLP-1 should ideally be validated by measuring in parallel plasma samples with high endogenous and exogenous levels of GLP-1, glucagon, and oxyntomodulin by a validated immunoassay and by the Olink proteomic assay.

Plasma gcg showed a positive association with BCAA. BCAA catabolism is not regulated by glucagon, and BCAA do not directly stimulate glucagon secretion [38, 39]. However, BCAA do play an important role in the pathogenesis of insulin resistance [42, 43] and the correlation between gcg and BCAA may reflect the mutual relationship between hepatic insulin and glucagon resistance [24]. A limitation of the current study is the lack of markers of insulin resistance, hereof plasma insulin levels. We previously observed that glucagon resistance and insulin resistance may coexist but importantly also occur independently of each other highlighting the differential pathophysiological mechanisms underlying glucagon and insulin resistance [28].

In agreement with previous studies, we found a link between MASLD and glucagon. In our analyses, we utilized MRI to estimate and diagnose MASLD based on hepatic steatosis, but we lacked data on the severity of fibrosis, which is the most critical predictor of clinical outcomes in MASLD. However, given the probable close relationship between glucagon and the metabolic alterations in MASLD, the degree of hepatic steatosis is likely to indicate the risk of developing cardiometabolic conditions, including diabetes. Our findings therefore support speculations that there is a two-way connection between MASLD and diabetes and suggest that glucagon could be a main factor in the pathogenesis.

Consistent with prior research, we identified an association between plasma gcg and individuals with type 2 diabetes at baseline. It is possible that this relationship may not only reflect the pathophysiological traits of diabetes but also the glucagonostatic effects of glucose-lowering drugs (e.g. metformin). In our analysis, however, we still detect a significantly increased plasma level of gcg in patients with type 2 diabetes.

Type 2 diabetes is often diagnosed by general practitioners, and only ∼41% of diabetes diagnoses are registered in hospital records [20]. Primary care data are linked to the UK Biobank for ∼45% of the participants up until 2016 (England) and 2017 (Scotland and Wales), so to get a longer follow-up period, incident type 2 diabetes was defined from secondary care ICD-10 diagnostic codes. This is a limitation of this study and may explain the flattening of the Kaplan-Meier curve (Fig. 2a) in the later years of the follow-up period, as hospital diagnoses may be registered at a later point than the primary care diagnosis. Another limitation is that quantification of liver fat (MRI PDFF) is obtained from the MRI scan performed ∼10.5 years after the baseline data was obtained.

Traditionally, the primary focus on glucagon receptor signalling has centred on the adenylate cyclase/cAMP/protein kinase A pathway as the predominant mediator of the hepatic glucose-mobilizing actions of glucagon. However, compelling evidence suggests that the PLC/IP3 pathway may be even more important for physiological levels of glucagon as compared to pathologically or pharmacologically elevated levels [44]. Further investigation into the impact of rare missense variants on this pathway is warranted.

The most common missense variant in the glucagon receptor, G40S has normal to mildly reduced cAMP signalling [45–47], and significantly decreased β-arrestin 1 signalling [8]. G40S has previously been linked to non-insulin-dependent diabetes and central adiposity in France and Sardinia [29, 30], but interestingly not in Japan or Finland [48, 49]. We did not find an increased prevalence of type 2 diabetes or obesity in heterozygotes or homozygotes of the G40S variant in the UK Biobank. However, there was a significant interaction between plasma gcg and G40S, with gcg having a significantly stronger association with BMI in carriers of G40S compared to wild-type controls (Tabel 2). On the contrary, the association between gcg and type 2 diabetes was borderline significantly lower in individuals with G40S compared to wild-type controls (Table 1). Together with the divergent literature on the effects of G40S on metabolic disorders, our results suggest that G40S may play a role in the development of type 2 diabetes and obesity, but that this may require parallel metabolic disruptions leading to altered gcg levels.

A tendency toward increased risk of obesity was previously shown for the group of cAMP LoF variants [8]. Since then, exome sequencing data was increased to cover the whole UK Biobank. After an approximately doubling of the sample size of carriers of a cAMP LoF variant, this tendency disappeared, both when BMI was treated as a continuous and dichotomized variable. However, the increased plasma levels of gcg in this group of individuals suggest that cAMP signalling is involved in the regulation of a factor, that in turn regulates gcg levels in a feedback manner. This has previously been suggested to be amino acids, with alanine being particularly important for this feedback system termed the liver-alpha cell axis [50–52]. In line with this, there was a tendency to increased alanine (p = 0.1) in individuals with cAMP variants adjusting for age, sex, BMI, fasting time, and creatinine.

The Frameshift category of glucagon receptors was defined rather broadly. Yet, carriers of one of the Frameshift variants were associated with significantly higher levels of liver fat compared to wild-type controls. This was not observed in G40S or the cAMP LoF variant groups, suggesting that a signalling pathway other than cAMP and β-arrestin is likely involved. Further functional subdivision and in silico predictions may help select and group variants that are pharmacologically characterized by more stratified loss-of-function phenotypes. Other research groups have found ubiquitination and β-arrestin to be essential for the signalling and internalization of the glucagon receptor [7, 53], whereas others report only a minor internalization of the glucagon receptor [54], suggesting that this pathway may not be a major regulator of glucagon receptor signalling. The multiple factors impacting signalling pathways are complex and warrant further exploration.

In conclusion, our study supports the involvement of glucagon signalling in metabolic disorders such as type 2 diabetes and MASLD and that increased gcg levels may predispose to type 2 diabetes. Furthermore, the presence of hyperglucagonemia in obesity, MASLD and type 2 diabetes indicates that distinct mechanisms may drive increased alpha cell secretion. Other signalling pathways than cAMP and β-arrestin recruitment may be important for the metabolic effects of glucagon such as the regulation of liver fat. Identification of the specific molecular characteristics responsible for the beneficial effects of glucagon on hepatic lipid turnover may be crucial in developing improved glucagon co-agonists for the treatment of MASLD and obesity.

## Abbreviations

Gcg: proglucagon
GLP-1: Glucagon-like peptide-1
LoF: loss-of-function
MASLD: metabolic dysfunction-associated steatotic liver disease
NPX: Normalized Protein eXpression
PDFF: Proton density fat-fraction
UKB: UK Biobank

## Acknowledgement

This research uses data provided by patients and collected by the NHS as part of their care and support. Copyright © (2023), NHS England. Re-used with the permission of UK Biobank. This research used data assets made available by National Safe Haven as part of the Data and Connectivity National Core Study, led by Health Data Research UK in partnership with the Office for National Statistics and funded by UK Research and Innovation. We acknowledge the UKB and the study participants. This research has been conducted using the UK Biobank Resource under Application Number 61785. We furthermore thank Dr. Stefan Stender, Rigshospitalet, Copenhagen, Denmark for fruitful discussions on genetic variants and analyses hereof.

## Data availability

All data used in this work can be acquired from the UK Biobank (https://www.ukbiobank.ac.uk/). All coding is available through https://github.com/nicwin98/UK-Biobank-GCG.

## Authors’ relationships and activities

NJWA has received research support and speaker fees from Mercodia, Novo Nordisk, Merck, MSD, and Boehringer Ingelheim. LLG has received research support and speaker fees from Novo Nordisk, Pfizer, Becton, Dickinson, Alexion, Gilead, Sobi, Alexion, Immunovia, Norgine, Pfizer, Astra Zeneca.

## Contribution statement

MWS and NJWA conceived the study idea. MWS, SLG, and MEO analysed exome sequencing data. MWS, AB, and MKJ performed survival analyses. MWS analysed proteomics data and performed association analyses. MWS and NJWA wrote the first draft of the manuscript, and all authors revised and accepted the final version of the manuscript.

