## Supplementary data for "Determinants of plasma levels of gcg and metabolic impact of glucagon receptor signalling – a UK Biobank study"

### ESM

**ESM Table 1:**

UK Biobank covariates used in the study.

| Covariables | Definitions | UKB data field |
|---|---|---|
| **Age** | Age when attended the assessment center at the initial assessment, truncated to a full year. | 21003 |
| **Sex** | Acquired from central registry at recruitment. | 31 |
| **Race** | Categorical variable, white and non-white. Indicates samples who self-identified as 'White British' at the initial touchscreen questionnaire and have very similar genetic ancestry based on a principal components analysis of the genotypes. | 22006 |
| **Body mass index (BMI)** | Weight/height$^2$, kg/m$^2$. BMI when attended the assessment center at the initial assessment. | 21001 |
| **Liver fat** | %. Proton density fat fraction measured by MRI scan at instance 2. | 40061 |
| **Date attending assessment center** | Date of attending assessment centre. | 53 |
| **Date of death** | Date of death. Acquired from central registry. | 40000 |
| **Date lost to follow up** | Date lost to follow-up. This data was last updated in May 2017. | 191 |
| **Sex chromosome aneuploidy** | Samples which were identified as putatively carrying sex chromosome configurations that are not either XX or XY | 22019 |
| **HbA1c** | Baseline HbA1c levels, mmol/mol | 30750 |
| **Glucose** | Baseline glucose measurement, mmol/L | 30740 |
| **Weekly alcohol consumption** | Calculated as the sum of beer, cider, champagne, white wine, red wine, fortified wine, spirits, and other alcoholic drinks. | 1588, 1578, 1608, 5364, 1568, 1598 |

| | | |
|---|---|---|
| **Amino acids** | Plasma levels of amino acids measured by NMR metabolomics | 23460, 23461, 23462, 23463, 23465, 23466, 23467 |
| **Proteomics** | Only available through the Research Analysis Platform. A list of field names are available at https://github.com/nicwin98/UK-Biobank-GCG | |

**ESM Table 2**

Mutations in the glucagon receptor included in the Frameshift variants group

| Frameshift mutations | | |
|---|---|---|
| Chrom_pos_ref_alt | N | Sequence Ontology |
| chr17_81809019_A_G | 1 | start lost |
| chr17_81809021_GC_G | 2 | frameshift truncation |
| chr17_81809027_CT_C | 1 | frameshift truncation |
| chr17_81809039_G_GCGACC | 1 | frameshift elongation |
| chr17_81809040_C_T | 5 | stop gained |
| chr17_81809045_CCT_C | 22 | frameshift truncation |
| chr17_81809064_C_CTGCT | 33 | frameshift elongation |
| chr17_81809838_C_A | 1 | stop gained |
| chr17_81810872_AC_A | 1 | frameshift truncation |
| chr17_81810888_CG_C | 3 | frameshift truncation |
| chr17_81810897_TCTC_T | 5 | inframe deletion |
| chr17_81810907_CT_C | 40 | frameshift truncation |
| chr17_81811122_TG_T | 1 | frameshift truncation |
| chr17_81811273_T_TACA | 1 | inframe insertion |
| chr17_81811283_CCCTGGGGGCCCTGCTCCTCGCCTTGGCCAT_C | 12 | inframe deletion |
| chr17_81811286_TG_T | 3 | frameshift truncation |
| chr17_81811293_C_GT | 1 | frameshift elongation |
| chr17_81811316_TG_T | 1 | frameshift truncation |
| chr17_81811316_T_TG | 14 | frameshift elongation |
| chr17_81811407_G_GCTGCA | 2 | frameshift elongation |
| chr17_81811422_CA_C | 1 | frameshift truncation |
| chr17_81811441_CTGTT_C | 1 | frameshift truncation |
| chr17_81811710_C_A | 6 | stop gained |
| chr17_81811751_T_TG | 1 | frameshift elongation |
| chr17_81811887_TGCCCC_T | 3 | frameshift truncation |
| chr17_81811911_CT_C | 1 | frameshift truncation |
| chr17_81811923_CAAGT_C | 2 | frameshift truncation |
| chr17_81812181_AG_A | 1 | frameshift truncation |
| chr17_81812216_G_A | 8 | stop gained |
| chr17_81812248_TC_T | 1 | frameshift truncation |

| | | |
|---|---|---|
| chr17_81812579_C_CA | 5 | frameshift elongation |
| chr17_81812581_ACTT_A | 47 | inframe deletion |
| chr17_81812598_CG_C | 1 | frameshift truncation |
| chr17_81812607_C_T | 2 | stop gained |
| chr17_81812838_CT_C | 1 | frameshift truncation |
| chr17_81812929_TC_T | 1 | frameshift truncation |
| chr17_81813477_C_T | 1 | stop gained |
| chr17_81813509_G_A | 3 | stop gained |
| chr17_81813529_G_A | 8 | stop gained |
| chr17_81813609_AG_A | 2 | frameshift truncation |
| chr17_81813624_C_T | 15 | stop gained |
| chr17_81813647_CT_C | 1 | frameshift truncation |

**ESM Table 3: Definition of diseases**

| | | |
|---|---|---|
| **Baseline type 2 diabetes** | Categorical variable. Cases were defined as probable and possible type 2 diabetes, and controls were defined as unlikely diabetes based on the Eastwood algorithm. | |
| **Incident type 2 diabetes** | Categorical variable. First, T2D was defined using ICD-10 codes for diabetes: E11 ("type 2 diabetes mellitus") and E14 ("unspecified diabetes mellitus") and the dates for the diagnosis (n = 3585). To refine the categorization further, the following steps were implemented:<br>1. Individuals additionally diagnosed with E10 ("type 1 diabetes mellitus") were removed from T2D (n = 360).<br>2. Individuals with probably and possibly type 2 diabetes based on the Estwood algorithm were removed from T2D (n = 1370).<br>3. Individuals with ICD10 code of T2D given prior to the baseline were removed from T2D (n = 304).<br>4. Individuals with baseline HbA1c > 48 mmol/mol were removed from T2D (n = 159).<br>This classification process resulted in 1,562 incident cases of T2D (893 men, 658 women). | 130708<br>130714<br>41270<br>30750 |
| **MASLD** | Categorical variable. Cases were defined as > 5.5% fat on MRI PDFF and an alcohol consumption < 30g/day for men and < 20g/day for women. Individuals with MRI PDFF > 5.5% and an alcohol consumption > 30g/day for men and > 20g/day for women were excluded from the control group. | 40061, 1588, 1578, 1608, 5364, 1568, 1598 |
| **Obesity** | Categorical variable. Cases were defined as BMI > 30 and controls as BMI < 25. | 21001 |

#### ESM Table 4

Baseline characteristics of the UK Biobank cohort and the sub-cohort included in the proteomics analysis. Prevalent type 2 diabetes (T2D) was defined from the Eastwood algorithm [1].

| Characteristic | UKB cohort | Proteomics sub-cohort |
|---|---|---|
| N | 408,931 | 40,158 |
| Male sex | 187,863 (45.9) | 18,585 (46.3) |
| Age | 56.9 (8) | 57.2 (8.1) |
| BMI (kg/m$^2$) [a] | 27.4 (4.8) | 27.4 (4.7) |
| Prevalent T2D | 17,719 (4.3) | 1842 (4.6) |
| Liver fat (%)*[a] | 4.8 (4.9) | 4.8 (4.8) |
| HbA1c (mmol/mol) [a] | 36 (6.5) | 36.1 (6.7) |

Continuous variables are presented as mean (SD) and categorical variables as n (%)

*Liver fat was quantified as MRI-PDFF at the second repeat (imaging) visit (median time from enrolment visit: 10.5 years).

[a] Missing data were present for BMI: a) n=1294 (0.3%), b) n=160 (0.4%); Liver fat: a) 384,507 (91.6%), b) 35,910 (89,4%); HbA1c: a) n=19,135 (4.7%), b) 1812 (4.5%).
